# Social cognition deficits and its biometric signatures in the behavioral variant of Alzheimer’s disease

**DOI:** 10.1101/2022.02.07.22270260

**Authors:** Ellen H. Singleton, Jay L.P. Fieldhouse, Jochum J. van ‘t Hooft, Marta Scarioni, Marie-Paule E. van Engelen, Sietske A.M. Sikkes, Casper de Boer, Diana Bocancea, Esther van den Berg, Philip Scheltens, Wiesje M. van der Flier, Janne M. Papma, Yolande A.L. Pijnenburg, Rik Ossenkoppele

## Abstract

The behavioral variant of Alzheimer’s disease (bvAD) is characterized by early and predominant behavioral changes, resembling the clinical profile of the behavioral variant of frontotemporal dementia (bvFTD). Social cognition deficits form hallmark features in bvFTD and altered biometric responses to socioemotional cues have been observed in bvFTD. However, little is known about social cognition and its biometric signature in bvAD. In this explorative study, we investigated all levels of social cognition (i.e., level-1: perception, level-2: interpretation and level-3: reasoning), using the Ekman 60 faces test (level-1), Interpersonal Reactivity Index (IRI) and empathy eliciting videos (level-2), the Social Norms Questionnaire (SNQ) and moral dilemmas (level-3), while measuring eyemovements and galvanic skin response (GSR). We compared 12 patients with bvAD with patients with bvFTD (n=14), typical AD (tAD, n=13) and controls (n=15), using ANCOVAs and post hoc testing, adjusting for age and sex. Regarding *perception*, bvAD (40.1±8.6) showed lower scores on the Ekman test compared to controls (50.1±4.6, p<0.001), and tAD (46.2±5.3, p=0.05) and higher scores compared to bvFTD (32.4±7.3, p=0.002). Eyetracking during the Ekman test revealed that groups did not differ in dwell time on the eyes (all p>0.05), but bvAD (18.7±9.5%) and bvFTD (19.4±14.3%) spent significantly less dwell time on the mouth when viewing the faces than controls (30.4±10.6%, p<0.05) and tAD (32.7±12.1%, p<0.01). Regarding *empathy*, bvAD (11.3±4.6) exhibited lower scores on the IRI *Perspective Taking* subscale compared with controls (15±3.4, p=0.02) and similar scores to bvFTD (8.7±5.6, p=0.19) and tAD (13.0±3.2, p=0.43). The GSR to empathy eliciting videos did not differ between groups (all p>0.05). Regarding *knowledge of social norms*, bvAD (16.0±1.6) and bvFTD (15.2±2.2) showed lower scores on the SNQ than tAD (17.8±2.1, both p<0.05) and controls (18.1±1.3, both p<0.01). Regarding *moral reasoning*, no differences among the groups were observed in responses to moral dilemmas (all p>0.05), while only bvFTD (0.9±1.1) showed a lower GSR during the personal condition compared with controls (3.2±3.3 peaks per minute, p=0.02). In conclusion, bvAD showed a similar though milder social cognition profile and a similar eyetracking signature compared with bvFTD and greater social cognition impairments and divergent eyemovement patterns compared with tAD. Our results suggest that bvAD and bvFTD show reduced attention to salient features during facial expression perception, potentially contributing to their emotion recognition deficits. These social cognition and biometric measures provide important insights into the basis of behavioral changes in bvAD, and might be valuable for its clinical diagnosis.

## Introduction

The behavioral variant of Alzheimer’s disease (bvAD) is a rare and atypical variant of Alzheimer’s disease (AD), characterized by early and predominant behavioral and personality changes with underlying AD pathology [1-3]. The clinical phenotype of bvAD overlaps substantially with that of the behavioral variant of frontotemporal dementia (bvFTD)[3]. bvFTD is characterized by a wide variety of social cognition deficits that are thought to underlie the behavioral disturbances [4]. Social cognition refers to all processes necessary for adequate social behavior, and can be classified into three levels, including perception of, interpretation of and reasoning about social cues [5, 6]. First, a prerequisite for adequate social behavior is that social cues such as facial expressions and gestures are sufficiently and accurately perceived (level 1) [5, 6]. Second, these elementary perceptions are then interpreted to extract the affective states of the other person and form the basis for empathy and “theory of mind” (level 2) [5, 6]. Third, this ability of perspective taking then serves as necessary input to guide social decision making, as well as complex interactions between social reasoning based on knowledge of social norms and moral reasoning (level 3) [5].

Deficits along all levels of social cognition have been described in bvFTD [7] and may contribute substantially to their behavioral and personality changes. Social cognition deficits have been postulated as a possible underlying mechanism contributing to the bvAD phenotype as well [8-11]. However, these reports are mainly based on case studies. Moreover, social cognition tests are prone to confounding by deficits in other cognitive domains such as memory or executive functioning that are likely to be impaired both in bvAD and bvFTD. To overcome this hurdle, biometric measures have been used to capture experiential aspects of social cognition in the context of bvFTD. For example, eyetracking and galvanic skin response measures may yield more direct and sensitive measurements of emotions and social behavior [12], as previous studies have captured emotional blunting using galvanic skin response [13, 14] and deficits in emotion recognition using eyetracking [15] in bvFTD. As such, these tools may help unveil primary physiological processes underlying social cognitive functioning in addition to providing earlier and objective measures to capture decline in social cognition.

Group studies of social cognition test scores in conjunction with biometric measures in bvAD are currently lacking, and therefore the overlap and differences compared with both bvFTD and typical AD are largely unknown. In this exploratory study, we examined social cognition across all levels, including emotion recognition, empathy, knowledge of social norms and moral reasoning in bvAD compared to bvFTD, typical AD and controls, in conjunction with eyetracking and galvanic skin response measurements. We hypothesized that bvAD would show intermediate social cognition performance compared with bvFTD and typical AD (i.e. worse than tAD and better than bvFTD), based on the similar yet milder behavioral profile in bvAD compared with bvFTD [16].

## Methods

### Participants

Between February 2020 and October 2021, we included 12 patients clinically diagnosed with bvAD from the Amsterdam Dementia Cohort (ADC, the Netherlands) [17]. All bvAD cases fulfilled >2/6 core clinical criteria for bvFTD [18] in combination with amyloid-beta positivity based on CSF or PET examinations (see Table S1). According to recently proposed research criteria, all bvAD cases met criteria for at least “possible bvAD” (i.e., clinical bvAD in combination with positive amyloid-beta biomarkers [3], see table S1 for an overview of bvFTD symptoms in bvAD cases). In the same period, we consecutively included patients with “typical AD” (defined as amyloid-beta positive patients with MCI or AD dementia with an amnestic-predominant presentation) and patients with probable bvFTD according to the Rascovsky criteria [18] and with a negative amyloid status. Cognitively normal (CN) controls consisted of individuals with subjective cognitive decline with CSF amyloid levels in the normal range in whom objective cognitive impairment was ruled out by neuropsychological assessments during dementia screening [19]. Figure S3 provides an overview of the inclusion flow for each diagnostic group. Age, sex and level of education was ascertained for all participants. Level of education was classified using the Verhage system ranging from 1 (no or little education) to 7 (highest academic degree) [20].

### Experimental procedures

Participants underwent social cogition testing at the Alzheimer Center Amsterdam in a room with consistent lighting conditions. The test protocol included all tasks and biometric measures described below and had a total duration of approximately on hour. While participants were tested, their caregiver filled out questionnaires in the waiting room. In case of controls, relatives filled out the questionnaires at home and returned them by mail.

### Social cognition measures

All tasks were presented in the iMotions platform (iMotions 8.0, iMotions A/S, Frederiksberg, Denmark) that integrates several biometric measures, enabling the investigation of eye movements and galvanic skin response simultaneously in each participant, while performing a social cognition test (Figure 1). For level 1, *perception*, the Ekman 60 faces test was used to measure emotion recognition [21]. Subjects were asked to identify which of the six basic emotions (i.e., angry, sad, happy, surprise, fear or disgust) is shown by the facial expression on each of the 60 images. The test consists of 60 items, with 10 different male and female faces each expressing the six basic emotions. Each correct item is awarded 1 point, with scores between 0-60 for the total score and 0-10 for individual emotions. During the Ekman test, eye movement patterns were recorded using the iMotions platform (see section below).

**Figure 1.**
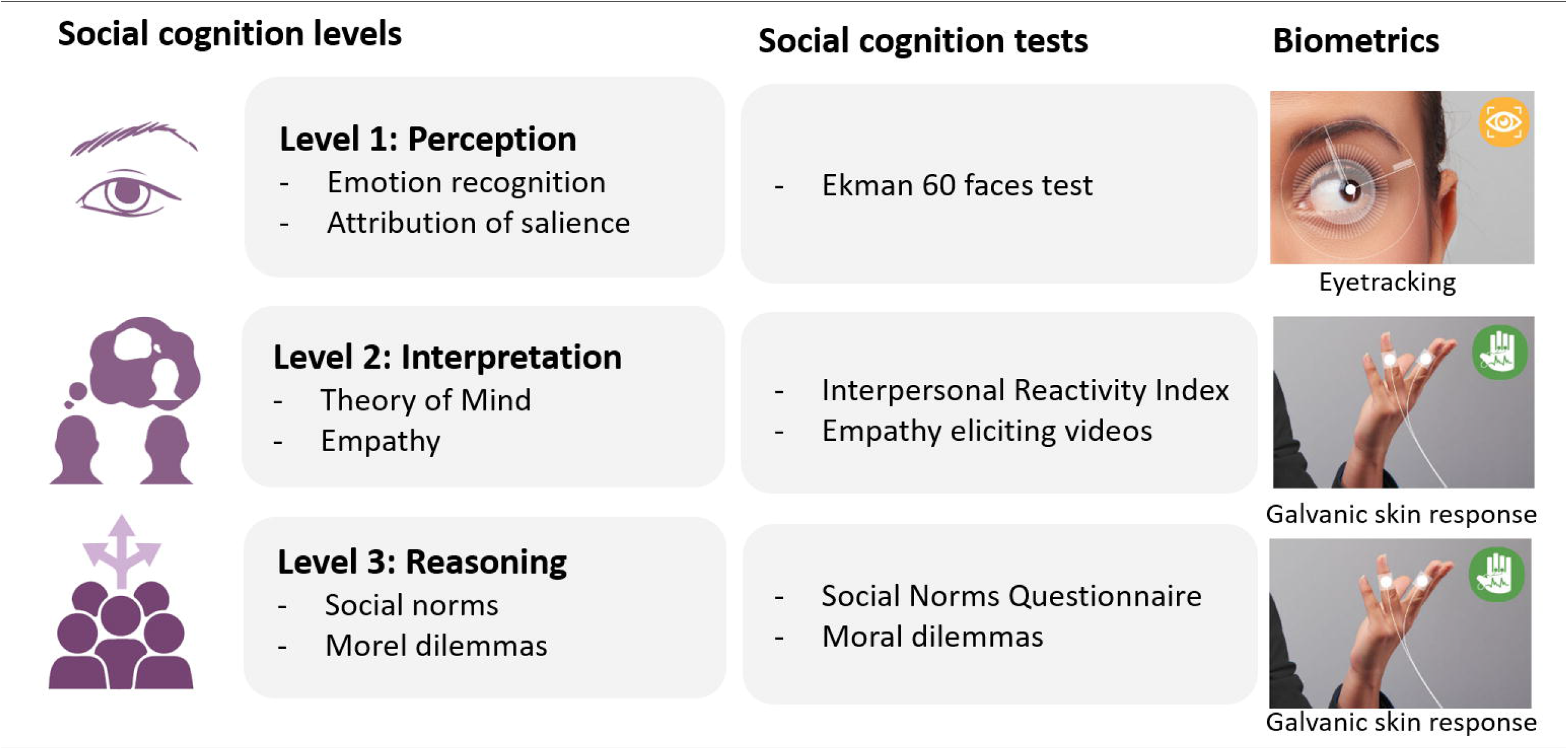
Social cognition framework and biometrics implemented in the current study

For each item, a 5 seconds window was presented within the platform with the face only, followed by the same face with the answer options below. There was no time restriction for participants to provide answers. For the eyetracking analyses, only data from the first 5 seconds window were included. Missing data were imputed using the R package “mice”, with 20% missing values for the Ekman as patients were able to progress without responding (see Figure S2 for a matrix of missing values per test). For level 2, *empathy*, two empathy eliciting videos were shown according to previously reported procedures [22]. Briefly, an “uplifting” empathy eliciting video about a surf project for children with autism and Down syndrome and a “distressing” empathy eliciting video about a charity foundation for severely malnourished children were shown. The duration of each video was between 60 to 80 seconds. Immediately after watching the video, participants were asked to rate on a 5-point Likert-scale (1=not at all, 5=extremely) the degree to which they felt “sympathetic”, “moved”, “compassionate”, “disturbed”, “upset”, and “worried”. The three former phrases were averaged as a measure of empathetic concern, while the average of the three latter phrases was used as a measure of personal distress [23, 24]. Galvanic skin response was recorded during this task with the iMotions platform (see below). In addition, the Dutch version of the Interpersonal Reactivity Index was administered in informants [25]. This is a questionnaire consisting of 28 items with a 5-point Likert scale, measuring 4 subscales: perspective taking (PT, the tendency to adopt another’s psychological perspective), empathetic concern (EC, the tendency to experience feelings of warmth, sympathy, and concern toward others), fantasy (FS, the tendency to identify strongly with fictitious characters), and personal distress (PD, the tendency to have feelings of discomfort of concern when witnessing other’s negative experiences). The percentage of rational answers was calculated per diagnostic group per condition. For level 3, *knowledge of social norms*, the Dutch version of the Social Norms Questionnaire was administered in participants, consisting of 22 questions assessing the ability to understand and identify social boundaries [26], each marked as correct or incorrect, with a total score between 0-22. A break score indicated a tendency to break social norms (for example indicating whether or not it is socially acceptable to tell a stranger you think he or she is overweight) and an overadherence score indicated a tendency to overadhere to a social norm (i.e., applying a social rule too rigidly, for example indicating that it is not socially acceptable to laugh when you trip and fall yourself). Missing data were imputed using the R package “mice” with 11% missing values for the SNQ test (see Figure S2 for a matrix of missing values per test). For *moral reasoning*, two classic moral dilemmas were presented to participants, consisting of the trolley (impersonal) and footbridge (personal) dilemma [27]. In the trolley dilemma participants were asked whether they would hit a switch that would redirect a trolley that is on its way to kill five individuals on a train rail, to a train rail heading for one individual (yes/no). In the footbridge dilemma, participants are asked whether they would push a man off a bridge in order to stop the train from killing the five individuals. These stories were presented using prerecorded audio fragments and were additionally presented in text on the screen. During this task, galvanic skin response was administered using the iMotions platform (see section below).

### Biometric measures

Eye movements were recorded with a Tobii Pro X2-60 screen-based eye tracker (Tobii, Stockholm, Sweden) with a sampling rate of 60 Hz. The tasks were presented on a 24” monitor with a screen resolution of 1920×1200 pixels. Patients were positioned between 55 and 75 cm from the screen. Before the task, participants completed a nine-point calibration procedure to ensure optimal eye tracking accuracy. The eyetracker uses near-infrared technologies to track and calculate gaze points. The dwell time is recorded as the percentage of time that the gaze was directed in a specific (manually-defined) area of interest during presentation of the stimulus. Areas of interest of the same size were drawn on the eyes and mouth of each Ekman face (see Figure S1 for an example) as these form the most salient features of the face to extract emotions [28]. Galvanic skin response was measured using the Shimmer 3 GSR+ system (Shimmer, Consensys, Dublin, Ireland, https://shimmersensing.com/) attached to the plantar side of two fingers of participants’ non-dominant hand. Signals were sampled at 128 Hz. Data were online band-pass filtered between 0.01 Hz and 1 Hz, and were subsequently analyzed through a standardized R notebook (see Table S2 for details) according to previously reported procedures [29], resulting in galvanic skin response peaks per minute per stimulus per respondent.

### Statistics

Differences in demographic variables were assessed using *X*^2^ tests for dichotomous data and ANOVAs for continuous variables. Differences among groups were assessed using ANCOVAs and emmeans post hoc tests, adjusting for age and sex, in R version 4.0.2. Supplemental data additionally shows the results without adjusting for age and sex (see Table S3-S9). Due to the exploratory nature of the study, no correction for multiple testing was applied.

## Results

### Demographic characteristics

Demographic characteristics are shown in Table 1. Patients with bvAD had an average age of 66.6±5.7 vs 64.6±6.7 in typical AD, 66.4±7.0 in bvFTD and 61.7±6.3 in CN. 75% of bvAD cases were male, versus 38.5% in typical AD, 64.3% in bvFTD and 37.5% in controls. No significant differences were found in MMSE scores or education between groups (all p>0.05) and bvAD (60.0%) and tAD (83.3%) showed higher proportions of APOE carriers than bvFTD (17.0%) and controls (33.3%, p=0.02).

**Table 1.** Demographic characteristics across diagnostic groups

|  | bvAD | tAD | bvFTD | Controls | p-value |
| --- | --- | --- | --- | --- | --- |
| N | 12 | 12 | 14 | 14 |  |
| Age | 66.6 (5.7) | 64.6 (6.7) | 66.4 (7.0) | 61.7 (6.3) | 0.09* |
| Sex, % male | 75 | 38.5 | 64.3 | 37.5 | 0.15** |
| MMSE | 24.8 (2.5) | 24.7 (4.6) | 26.2 (2.3) | 28.1 (1.3) | 0.08* |
| Education<br>(Verhage score 1-7) | 5.1 (1.4) | 5.8 (1.3) | 5.4 (0.9) | 5.7 (0.7) | 0.29* |
| APOEε4, % carrier | 6/10 (60.0%) | 10/12 (83.3%) | 1/6 (17.0%) | 4/12 (33.3%) | **0.02\*** |

Data are presented as mean(SD); n (%); *bvAD*=behavioral variant of Alzheimer's disease, *tAD*=typical Alzheimer's disease, *bvFTD*=behavioral variant of frontotemporal dementia, *MMSE*=mini mental state examination.
\*Based on an ANOVA test
\*\*Based on $\chi^2$ test

### Level 1: Perception

#### Ekman 60 faces test scores and eyetracking

Participants with bvAD (40.1±8.6) showed lower scores on the Ekman 60 faces test compared with controls (50.1±4.6, p<0.001), and typical AD (46.2±5.3, p=0.05) and higher scores compared to bvFTD (32.4±7.3, p=0.002, Figure 2). Compared with typical AD, bvAD showed lower scores on the angry faces (6.2±2.4 vs 8.4±1.3, p=0.02) and did not differ on the other emotions (all p>0.05). bvAD and bvFTD did not differ on any emotion (all p>0.05). Compared to controls, bvAD showed significantly lower scores on the angry (6.2±2.4 vs 8.9±1.4, p=0.006), disgusted (5.4±2.5 vs 8.4±1.6, p=0.0004) and surprised (8.1±1.7 vs 9.1±0.7, p=0.02) faces and did not differ on the other emotions (all p>0.05). Despite comparable dwell time in the eyes across groups (all p>0.05; Table S4), bvAD (18.7±9.5) showed lower dwell time percentages in the mouth area-of-interest compared to controls (30.4±10.6, p=0.008) and typical AD (32.7±12.1, p=0.004), and did not differ from bvFTD (19.4±14.3, p=0.78). This pattern was observed across all six emotions (Figure 3 and Table S5).

**Figure 2.**
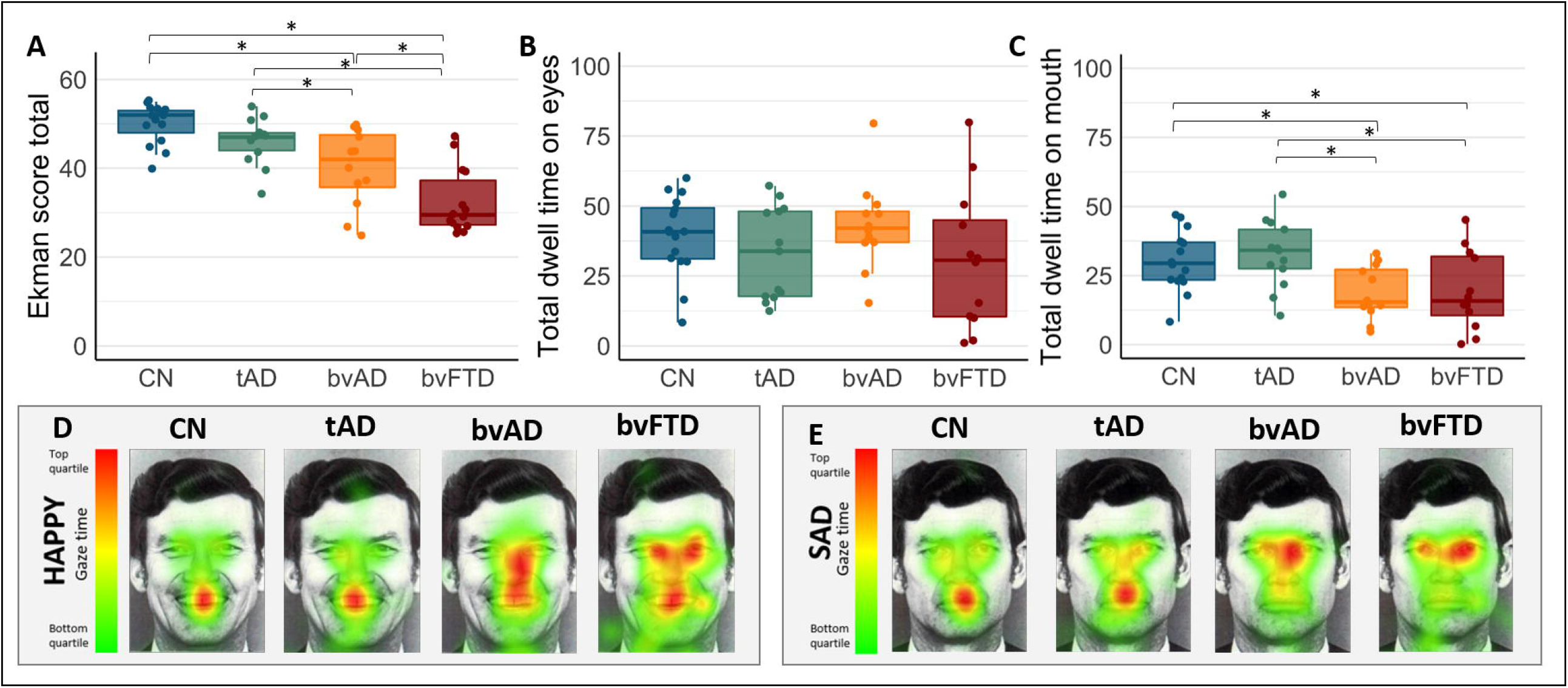
Perception (level 1): Emotion recognition measured by the Ekman 60 faces test scores and total eyetracking dwell time across groups The dwell time is a percentage of time that participants gaze was upon certain features of the image of the total time the stimulus was presented. The pixels with the highest amount of time spent by gaze are represented by red colors while the pixels with the lowest amount of time spent by gaze are represented by green colors. *CN*=cognitively normal controls, *tAD*= typical AD, *bvAD*=behavioral variant of AD, *bvFTD*= behavioral variant of frontotemporal dementia.

**Figure 3.**
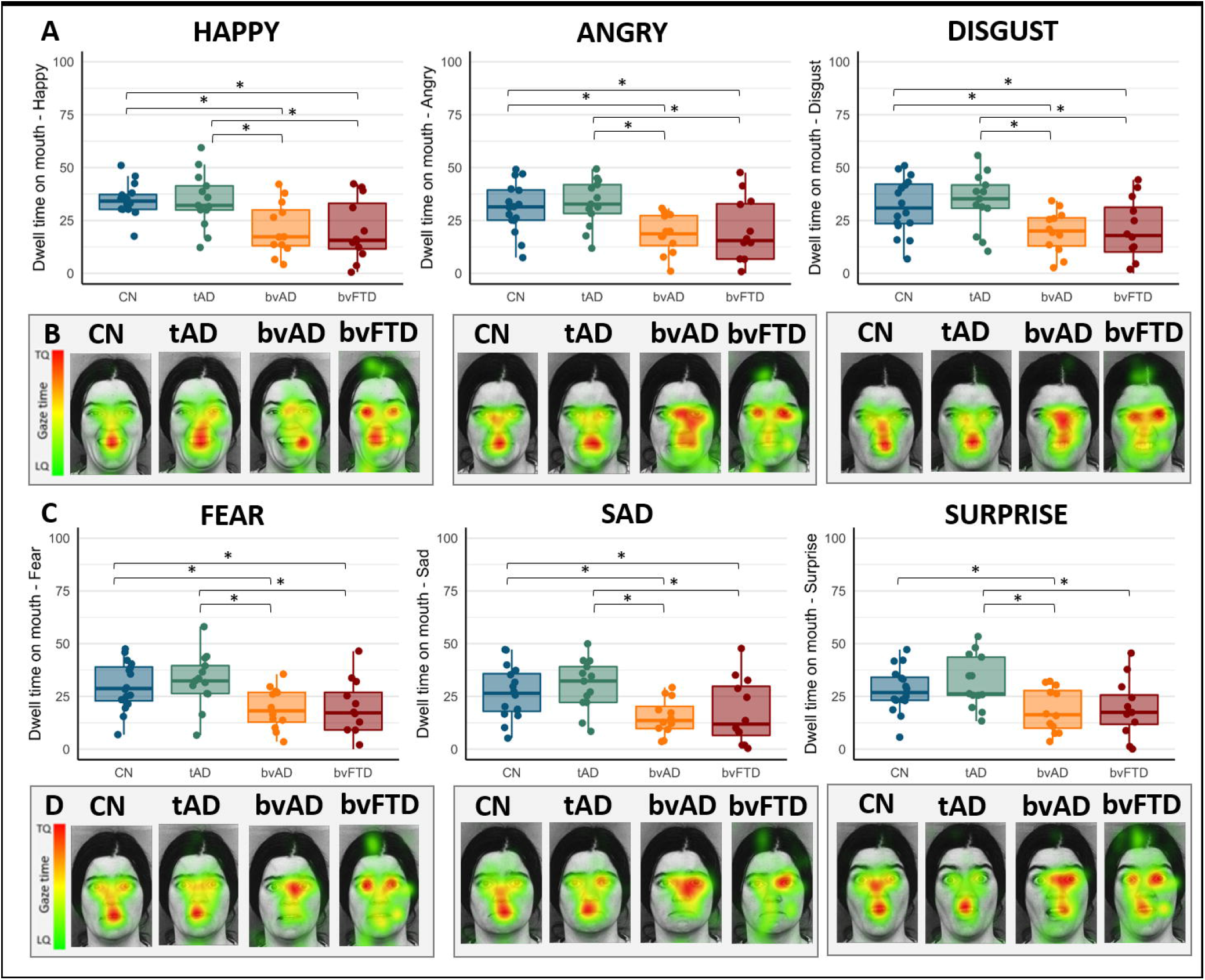
Results of Perception (level 1): heat maps of eyetracking dwell time on the mouth in Ekman 60 faces test images across groups The dwell time is a percentage of time that participants gaze was upon certain features of the image of the total time the stimulus was presented. The pixels with the highest amount of time spent by gaze are represented by red colors while the pixels with the lowest amount of time spent by gaze are represented by green colors. *CN*=cognitively normal controls, *tAD*=typical AD, *bvAD*=behavioral variant of AD, *bvFTD*=behavioral variant of frontotemporal dementia, *TQ*=top quartile, *LQ*=lower quartile.

### Level 2: Interpretation

#### Interpersonal Reactivity Index

On this informant-rated questionnaire measuring empathy, bvAD showed lower scores on the Perspective Taking subscale of the Interpersonal Reactivity Index (10.1±3.0 vs 15.5±3.4, p=0.04) and Fantasy subscale (10.3±5.2 vs 13.6±1.8, p=0.008) compared to controls (Figure 6 and Table S6), while showing no significant differences with typical AD (Perspective Taking: 13.0±3.2, Fantasy subscale: 13.0±3.1) and bvFTD (Perspective Taking: 8.7±5.6, Fantasy subscale: 13.6±1.8, all p>0.05). No significant differences were observed between groups on the Empathetic Concern and Personal Distress subscales (all p>0.05) of the Interpersonal Reactivity Index (Figure 4 and Table S6).

**Figure 4.**
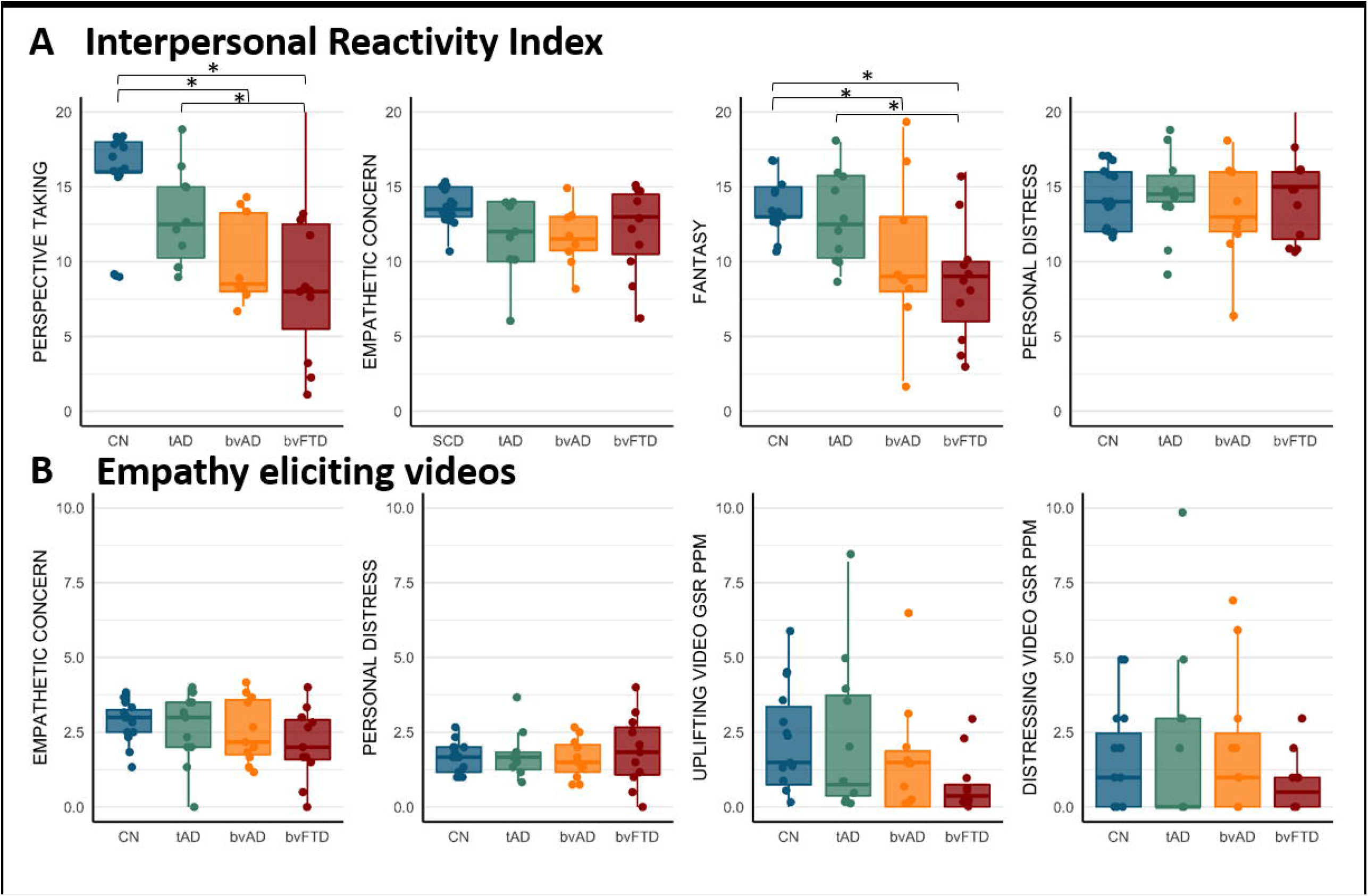
Results of Interpretation (level 2): Interpersonal Reactivity Index scores and responses to empathy eliciting videos across groups *GSR*=galvanic skin response, *PPM*=peaks per minute, *CN*=cognitively normal controls, *tAD*= typical AD, *bvAD*=behavioral variant of AD, *bvFTD*=behavioral variant of frontotemporal dementia.

#### Empathy eliciting videos

No significant differences were found among groups in empathetic concern and personal distress scores after watching empathy eliciting videos, nor in their galvanic skin response while watching those videos (Figure 4 and Table S7).

### Level 3: Reasoning

#### Knowledge of social norms

bvAD (16.0±1.6) showed lower scores on the Social Norms Questionnaire total score compared to controls (18.1±1.3, p=0.02), and typical AD (17.8±2.1, p=0.04; Figure 5 and Table S8). No significant differences were found among groups on the break score, while only bvFTD (5.1±2.3) showed a higher overadherence score compared to controls (2.5±1.1, p=0.001) and typical AD (3.1±2.1, p=0.01). Analyses without adjusting for age and sex indicated that bvAD showed a higher overadherence score compared to controls (3.1±2.1 vs 2.5±1.1, p=0.05), suggesting a possible tendency to apply social norms too rigidly.

**Figure 5.**
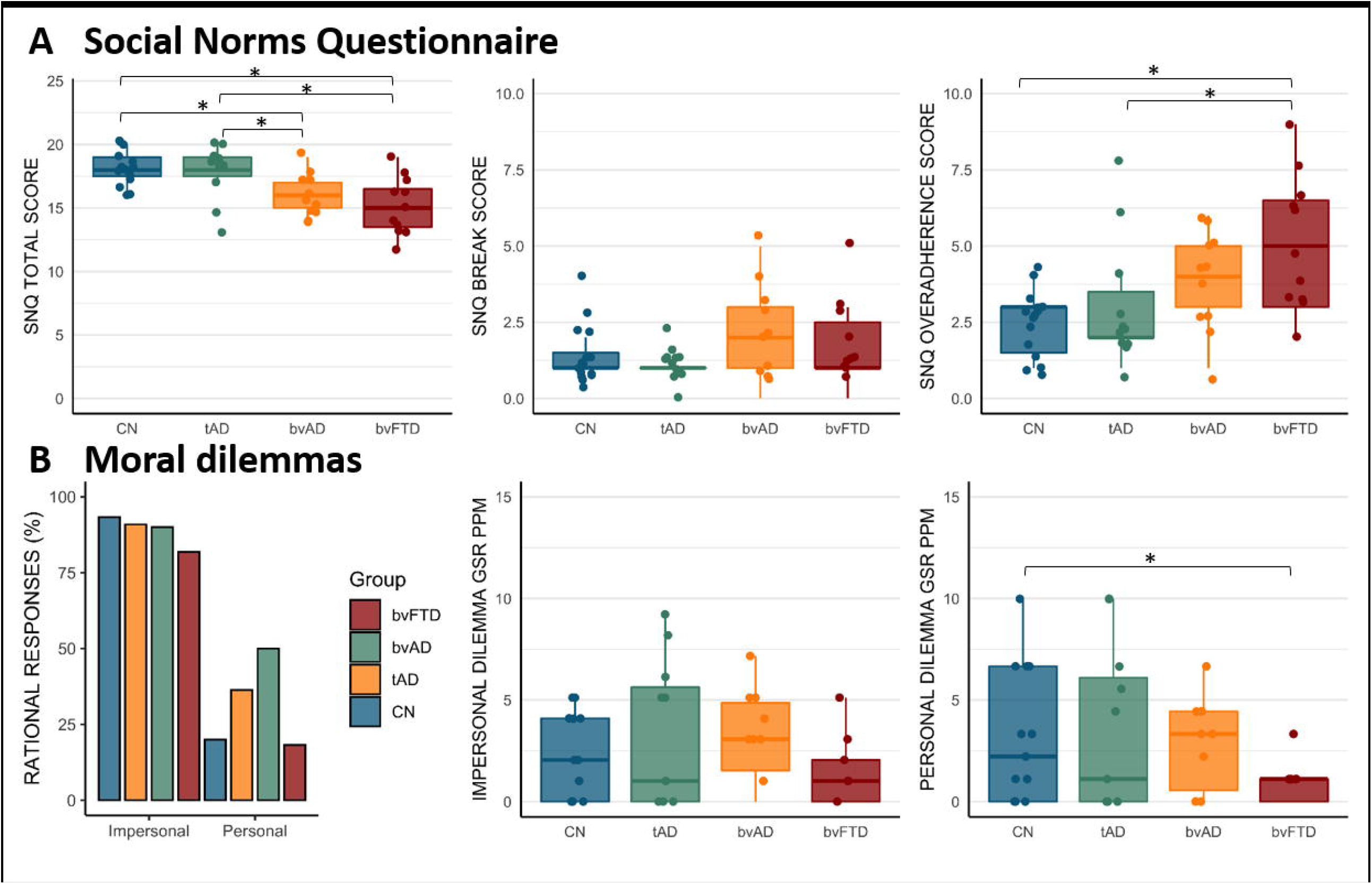
Results of Reasoning (level 3): knowledge of social norms and moral reasoning as measured by the Social Norms Questionnaire and (galvanic skin response to) moral dilemmas across groups *GSR*=galvanic skin response, *PPM*=peaks per minute, *CN*=cognitively normal controls, *tAD*=typical AD, *bvAD*=behavioral variant of AD, *bvFTD*=behavioral variant of frontotemporal dementia.

#### Moral reasoning

No significant differences were found among groups in the percentage rational responses provided to moral dilemmas. Except for lower galvanic skin response peaks per minute in the personal dilemma condition in bvFTD compared to controls (0.9±1.1 vs 3.2±3.3, p=0.02; Figure 7 and Table S9), there were no differences in galvanic skin response peaks per minute to moral dilemmas across groups (2.9±2.3 in bvAD vs 3.4±4.1 in tAD and 3.2±3.3 in controls, all p>0.05; Figure 7 and Table S9).

## Discussion

In this exploratory study, we found social cognition deficits on all three levels of social cognition in bvAD and a biometric signature that overlapped with bvFTD in level 1. The impairments on emotion recognition in patients with bvAD were observed in parallel with lower dwell time on the mouth while viewing emotional facial expressions compared to controls and typical AD. On the second level, deficits in empathy were reported by bvAD informants, while no differences were observed on subjective and biometric responses to empathy eliciting videos. On the third level, bvAD patients showed impairments in knowledge of social norms. Analyses without covariates indicated higher overadherence scores in bvAD than controls, suggesting a tendency to apply social rules too rigidly. No differences were observed in the subjective and biometric responses to moral dilemmas across diagnostic groups. This comprehensive study of social cognition in bvAD, combining a wide range of social cognition tests with biometric measures, points towards deficits across all levels of social cognition, showing a similar yet milder profile compared to bvFTD, and distinct elementary perceptual processes in bvAD compared to typical AD.

A main finding in the present study was that eye movement patterns showed lower dwell time on the mouth and similar dwell times on the eyes in bvAD and bvFTD compared to typical AD and controls. This differs from eye movement patterns observed in other conditions where social cognition is altered. For example, reduced social cognition in autism is observed in parallel with a lack of gaze on the eyes [30], while Williams syndrome (a hypersocial developmental disorder) is characterized by hyperfixation on the eyes [31]. Our results also differ from recent work in bvFTD showing increased fixations on the eyes while spending the same amount of time on the mouth as controls [28, 32]. A commonality between all the studies is the suggestion of a mechanism by which patients with bvFTD “look but don’t see”, as they do spend most time on the salient features of the face. The question whether this represents a deficit in *encoding* or *interpretation* of emotionally salient stimuli is a topic of debate. The fact that the previous study showed a hyperfixation on the eyes in bvFTD may point towards *interpretation* deficits or compensatory mechanisms, while our findings of lower dwell time on the mouth in bvAD and bvFTD may suggest that the elementary perceptual processes (i.e. *encoding*) are altered in these diseases, as they may not utilize all relevant facial features to extract emotional meaning. In addition, patients may spend more time on other features of the face than the eyes and mouth that hold less relevance for accurate detection of emotions. Either way, our results suggest that the analysis of eyetracking patterns may hold strong potential for the differentiation of bvAD and bvFTD from typical AD and controls. As such, eyetracking may form a valuable tool for the early and objective detection of social cognition alterations in these disease entities, either by detecting early changes in socioemotional functioning when social cognition scores may not be impaired yet or by bypassing cognitive impairments in advances cases.

Despite evident differences in the perceptual level of social cognition in bvAD, these patients did not exhibit lower scores on all tests across all levels of social cognition compared with controls and typical AD. For example, they did not show differences in their own emotional valuations after watching empathy eliciting videos or the amount of rational responses to moral dilemmas, nor in galvanic skin response to those tasks. Regarding moral dilemmas, this may be due to the high cognitive demand of the tasks, hampering patients’ understanding of the dilemmas, while the empathy eliciting videos draw patients’ attention exogenously and require less cognitive engagement. Indeed, in the uplifting empathy eliciting video, a trend towards lower galvanic skin response was observed in bvAD compared to controls, and bvFTD showed a significantly lower galvanic skin response compared to controls (Figure 4 and Table S7). In addition, while a clinically well-validated test was used on level 1 (i.e. the Ekman 60 faces test), more experimental tests were used on other levels (i.e. empathy eliciting videos and moral dilemmas), with lower variance in scores. Furthermore, the duration of the test protocol was rather long (∼1 hour) and fatigue may have influenced results especially towards the end of the session (i.e. moral dilemmas). This may have hampered finding group differences in these small samples. In clinical practice, the administration of one test of social cognition (i.e. the Ekman test at level 1) may be sufficient to detect differences, while a full screening may be beneficial for adequate psychosocial interventions aimed at a comprehensive understanding of disease mechanisms.

Our results expand upon the scarce existing literature on social cognition deficits in bvAD, which is mainly based on case studies. Emotion recognition deficits were reported in a sample of eight bvAD cases using the mini-SEA (Social cognition & Emotional Assessment) [8], and two case studies using the Facial Emotion Recognition Test and the emotion recognition subtests of the TASIT (The Awareness of Social Inference Test) [9, 11]. These studies involved patients with an initial bvFTD diagnosis, who had an AD biomarker profile. Impairments of Theory of Mind (ToM) were reported in the sample of eight bvAD cases using the mini-SEA[8] and one case study describing bvAD using the Reading the Mind in the Eyes Test and the TOM-15[10]. Impairments in knowledge of social norms were reported in one clinical bvFTD case with AD biomarkers previously based on the Social Norm Knowledge Questionnaire [10]. The limited studies on social cognition in bvAD showed a lack of inclusion of tests along all levels of social cognition, and a lack of measurements capturing the experiential, perceptual and non-cognitive processes of social cognition. Our findings in a group study including tests along levels of social cognition firmly established deficits in emotion recognition, empathy and knowledge of social norms in bvAD. In addition, the combination of social cognition tests and biometric measurements yielded insights into the elementary perception of emotional facial expressions, showing differential primary processing in bvAD and bvFTD compared with controls and typical AD. The elementary processing of social cues may contribute to higher order social cognition deficits in bvAD and bvFTD. Although social cognition deficits were less profound in bvAD than in bvFTD in the current study, it remains to be elucidated in larger samples if the profile of social cognitive deficits in bvAD is similar to or milder than that of bvFTD. Nevertheless, our results suggest that specific social cognition tests and biometrics may be valuable in the clinical diagnosis of bvAD, such as the Ekman 60 faces test, Interpersonal Reactivity Index and Social Norms Questionnaire and eye movement patterns. The introduction of the first set of research criteria for bvAD [16] may greatly improve the general recognition of bvAD and the addition of the aforementioned tools may increase the diagnostic accuracy of bvAD even further in the future.

The neurobiological origins of the observed social cognition deficits in bvAD are poorly understood. Neuroimaging studies have shown either a mix of anterior and posterior predominant pattern or a predominant temporoparietal pattern of neurodegeneration based on atrophy or hypometabolism [2, 33-35], with a relative lack of involvement of anterior brain regions. Compared with bvFTD, bvAD showed less involvement of the salience network, which is a network that regulates socioemotional processing and social cognition [36]. Moreover, neuropathological small samples of bvAD patients suggested that they may not show a selective loss of Von Economo Neurons (VENs) in the anterior cingulate cortex [37, 38], which are specialized neurons located within key regions of the salience network that sub serve social functioning in humans and highly intelligent mammals [39]. Therefore, traditional regions implicated in social cognition in bvFTD may not underlie social cognition deficits in bvAD. In typical (amnestic-predominant) AD, different regions have been proposed to underlie social cognition, including the precuneus, posterior cingulate cortex, temporoparietal junction and hippocampus, either directly or indirectly [4]. Alternatively, unique “bvAD” features such as anterior default mode network involvement [40] or altered amygdalar volumes [40] compared to typical AD may contribute to the observed deficits in bvAD, in addition to mild involvement of “bvFTD”-specific regions, such as subtle frontoinsular involvement [40].

The strength of this study is that we applied a comprehensive social cognition test battery spanning all levels of social cognition and combined it with biometric measurements in biomarker-confirmed bvAD patients. There are also limitations. First, although this is the largest study of its kind to date, the sample size of this exploratory study was modest. Since galvanic skin response can show substantial variation across participants, the small samples sizes may have hampered the detection of group differences. Second, the groups differed substantially in proportions of males vs females due to inherent overrepresentation of males in both bvAD [2] and bvFTD [41]. Since females are known to show better performance on social cognition tests [42], this may have influenced results in favor of the tAD and CN groups. However, as all results were corrected for age and sex the effects in the current work are deemed minimal. Third, due to the exploratory nature of this study and the low sample sizes, no correction for multiple testing was applied. Future, hypothesis driven, work with larger groups should incorporate adequate correction.

Future research should focus on multiple issues. Our findings should be replicated in larger cohorts, both in terms of social cognition tests and biometrics. Regarding the eyetracking, future work should especially investigate the role of encoding of the mouth as a salient facial feature for emotion recognition in the context of normal aging and disease, as well as investigating gaze patterns in more detail in bvAD and bvFTD. Moreover, future research should employ a similar study design with lower cognitive demands that sufficiently stimulates arousal to investigate the role of galvanic skin response to emotional stimuli. Future research should also incorporate more clinically validated tests for level 2 and 3 with larger statistical variety, which are suitable for the acquisition of biometric measurements in order to capture experiential processing in patients directly. In addition, future work should investigate the relationship between biometric measures and social cognition scores and their potential additive diagnostic accuracy above social cognition tests alone. Furthermore, future studies should investigate longitudinal patterns of changes in biometrics relative to social cognition test score changes and the added value of biometrics along different disease stages, in order to assess whether biometrics may indeed be more sensitive measures than social cognition test scores. Lastly, given that the neurobiological underpinnings of social cognitive deficits likely differ between bvAD and bvFTD, extensive profiling of social cognitive deficits and their biometric correlates in both disorders should be compared.

In conclusion, bvAD showed a similar though slightly milder pattern of social cognition deficits as observed in bvFTD, characterized by deficits in emotion recognition, empathy, and knowledge of social norms, with a similar eyetracking signature to bvFTD. Compared to typical AD, bvAD showed social cognition impairments in emotion recognition and knowledge of social norms and divergent eye movement patterns. Future research should focus on including larger sample sizes when assessing social cognition in conjunction with biometrics, incorporating social cognition tasks with low cognitive demands in bvAD, and assessing social cognition along all levels in bvAD, in comparison with bvFTD. These social cognition and biometric measures provide important insights into the basis of the behavioral and personality changes in bvAD, and might serve as valuable tools for an accurate diagnosis of bvAD in investigational and clinical settings in the future.

## Data Availability

Data is available upon reasonable request.

## Acknowledgements

Research of the Alzheimer center Amsterdam is part of the neurodegeneration research program of Amsterdam Neuroscience. The Alzheimer Center Amsterdam is supported by Stichting Alzheimer Nederland and Stichting VUmc fonds. WF holds the Pasman chair. The authors would furthermore like to acknowledge Chrissy Rijkers for her help in constructing the test protocol for the current manuscript, dr. Stefan van der Stigchel for his advice in the setup of the eyetracking part of the study, dr. Joke Spikman for her advice on the construction of the social cognition test battery, Mardou Leeuwestijn-Koopmans for her help in patient recruitment, and Kiara Heide and Oscar Haven for their support in constructing and monitoring the study test platform and conduction.

## List of abbreviations

Aβ: β amyloid
bvAD: behavioral variant of Alzheimer’s disease
tAD: typical Alzheimer’s disease
bvFTD: behavioral variant frontotemporal dementia
MCI: mild cognitive impairment
MMSE: mini mental state examination
APOE: Apolipoproteine E
ADC: Amsterdam Dementia Cohort
CSF: cerebrospinal fluid
PET: positron emission tomography
MRI: magnetic resonance imaging
IRI: Interpersonal Reactivity Index
SNQ: Social Norms Questionnaire
GSR PPM: Galvanic Skin Response Peaks Per Minute
AOI: Area of Interest

