## Supplemental data for "Social cognition deficits and its biometric signatures in the behavioral variant of Alzheimer’s disease"

**Table S1.** bvFTD core clinical symptoms and amyloid levels in bvAD cases

|  | | **bvAD_1_** | **bvAD_2_** | **bvAD_3_** | **bvAD_4_** | **bvAD_5_** | **bvAD_6_** | **bvAD_7_** | **bvAD_8_** | **bvAD_9_** | **bvAD_10_** | **bvAD_11_** | **bvAD_12_** |
| --- | --- | --- | --- | --- | --- | --- | --- | --- | --- | --- | --- | --- | --- |
| Sex | | M | M | M | M | M | M | F | F | M | F | M | M |
| Age | | 60-65 | 65-70 | 75-80 | 60-65 | 70-75 | 55-60 | 70-75 | 65-70 | 60-65 | 65-70 | 60-65 | 60-65 |
| MMSE | | 26 | 28 | 25 | 26 | 27 | 23 | 22 | 27 | 26 | 19 | 27 | 25 |
| bvFTD criteria | |  |  |  |  |  |  |  |  |  |  |  |  |
|  | Disinhibition | Y | Y | Y |  |  | Y | Y | Y |  | Y | Y | Y |
|  | Loss of empathy | Y |  |  | Y | Y | Y |  |  | Y |  |  |  |
|  | Compulsivity | Y | Y | Y |  | Y |  | Y | Y | Y |  |  | Y |
|  | Hyperorality | Y |  |  |  | Y |  |  | Y | Y |  |  |  |
|  | Apathy |  | Y |  | Y | Y |  |  |  | Y | Y | Y |  |
|  | Other |  | Y, delusions | Y, depression, delusions | Y, restlessness |  | Y, anxiety, depression | Y, delusions |  | Y, delusions |  |  |  |
|  | Total bvFTD symptoms | 4 | 3 | 2 | 2 | 4 | 2 | 2 | 3 | 4 | 2 | 2 | 2 |
| Amyloid confirmation (CSF/amyloid PET) | | Positive PiB-PET | A+/T+ CSF | A+/T+ CSF | A+/T+ CSF | A+/T+ CSF | A+/T+ CSF | A+/T+ CSF | A+/T+ CSF | Positive PiB-PET | A+/T+ CSF | A+/T+ CSF | Positive PiB-PET |

**Table S2**. Details on the galvanic skin response methodology used in the iMotions R notebook to extract peaks per minute

| 1 | The calibrated GSR signal from the sensor (Shimmer or BIOPAC EDA100C) is retrieved for the given stimulus and respondent (expressed in microSiemens, µS). |
| --- | --- |
| 2 | The sample rate is determined. Parameter [Gap interpolation length threshold [ms]] is used to identify possible gaps in the signal due to signal drops. If a gap in the signal is shorter than this threshold, then the missing data is linearly interpolated. Otherwise, the gap is left and each of the signal fragments is processed separately. Option [Remove signal discountinuities caused by the sensor switching range setting] should only be checked when the recorded signal has many discountinuities caused by the auto-range switching setting of Shimmer. |
| 3 | The phasic signal is extracted using a median filter over a fixed length time window: parameter [Phasic filter length [ms]]. The phasic signal is calculated by subtracting the running median from the calibrated signal, smoothing the ends of the signal by using symmetrical medians of subsequently smaller bandwidth, but for the very first and last value where Tukey's robust end-point rule is applied. |
| 4 | A low-pass Butterworth filter with cutoff frequency: parameter [Lowpass filter cutoff frequency [Hz]] is applied to the phasic signal (in order to remove powerline noise). |
| 5 | Onsets and offsets are detected on the phasic signal. Onsets are all the points where the phasic signal crosses above the onset threshold: parameter [Peak onset threshold [microSiemens]]. Offsets are the points where the phasic signal crosses below the offset threshold: parameter [Peak offset threshold [microSiemens]]. Each onset-offset pair defines a window, and the maximum value attained by the calibrated signal in a window is marked as a possible peak. The amplitude of a possible peak is defined as the difference between the maximum value of the calibrated signal in the window and its value at the onset point. A possible peak is selected if:  (1) Its amplitude is above the amplitude threshold: parameter [Peak amplitude threshold [microSiemens]], and  (2) the time difference between onset and offset is above the duration threshold: parameter [Minimum peak duration [ms]].  The selected peaks are returned. |

**Table S3.** Ekman scores across groups s

|  | bvAD | tAD | bvFTD | CN | p-value* | Group differences, unadjusted | Group differences, adjusted** |
| --- | --- | --- | --- | --- | --- | --- | --- |
| N_ekman_ | 12 | 13 | 14 | 15 |  |  |  |
| Happy | 9.50 (1.00) | 9.62 (0.96) | 9.43 (0.85) | 10.00 (0.00) | 0.07 | Ns | Ns |
| Angry | 6.17 (2.41) | 8.38 (1.33) | 4.79 (3.14) | 8.87 (1.41) | 0.0007 | bvAD<CN, p=0.003, bvFTD<CN, p<0.00001,  bvAD<tAD, p=0.01,  bvFTD<tAD, p<0.0000 | bvAD<CN, p=0.006, bvFTD<CN, p<0.0001, tAD<CN, p=0.02,  bvAD<tAD, p=0.02 bvFTD<tAD, p<0.0001 |
| Disgust | 5.42 (2.54) | 6.85 (1.68) | 3.71 (2.61) | 8.40 (1.59) | <0.00001 | bvAD<CN, p=0.0007, bvFTD<CN, p<0.00001, bvFTD<bvAD, p=0.05,  bvFTD<tAD, p=0.0004 | bvAD<CN, p=0.0004,  tAD<CN, p=0.02,  bvFTD<CN, p<0.0001,  bvAD>bvFTD, p=0.03 |
| Fear | 5.08 (2.78) | 6.15 (1.99) | 3.50 (2.35) | 6.60 (2.41) | 0.005 | bvFTD<CN, p=0.001, bvFTD<tAD, p=0.006 | bvFTD<CN, p=0.004, bvFTD<bvAD, p=0.07, bvFTD<tAD, p=0.01 |
| Sad | 5.83 (2.59) | 6.38 (2.26) | 3.71 (2.46) | 7.13 (2.03) | 0.0007 | bvFTD<CN, p=0.0002, bvFTD<bvAD, p=0.03,  bvFTD<tAD, p=0.004 | bvFTD<CN, p=0.0005,  bvFTD<bvAD, p=0.01, bvFTD<tAD, p=0.01 |
| Surprise | 8.08 (1.68) | 8.85 (1.28) | 7.29 (1.86) | 9.13 (0.74) | 0.008 | bvFTD<CN, p=0.001, bvFTD<tAD, p=0.007 | bvAD<CN, p=0.02, bvFTD<CN, p=0.0002,  bvFTD<tAD, p=0.002 |
| Total | 40.08 (8.63) | 46.23 (5.31) | 32.43 (7.26) | 50.13 (4.56) | <0.00001 | bvAD<CN, p=0.0002, bvFTD<CN, p<0.0001, bvAD<tAD, p=0.02,  bvFTD<bvAD, p=0.004, bvFTD<tAD, p<0.0001 | bvAD<CN, p=0.0008, bvFTD<CN, p<0.0001, bvAD<tAD, p=0.05,  bvAD>bvFTD, p=0.002,  tAD>bvFTD, p<0.0001 |

*based on anova test
**based on post hoc group comparisons
***based on post hoc group comparisons adjusted for age and sex

**Table S4.** Dwell time within Areas of Interest (AOI) Eyes in Ekman faces across groups s

|  | bvAD | tAD | bvFTD | CN | p-value* | Group differences, unadjusted | Group differences, adjusted ** |
| --- | --- | --- | --- | --- | --- | --- | --- |
| N_AOI_ | 12 | 13 | 12 | 16 |  |  |  |
| Happy | 30.99 (13.58) | 27.27 (13.58) | 27.27 (17.15) | 30.58 (12.47) | Ns | Ns | Ns |
| Angry | 43.74 (17.42) | 34.45 (16.03) | 32.50 (26.08) | 40.52 (17.42) | Ns | Ns | Ns |
| Disgust | 44.59 (17.62) | 33.37 (19.74) | 31.08 (25.79) | 39.86 (15.31) | Ns | Ns | Ns |
| Fear | 45.12 (17.68) | 45.12 (16.82) | 33.34 (26.26) | 42.52 (16.46) | Ns | Ns | Ns |
| Sad | 50.21 (17.62) | 38.11 (18.58) | 35.09 (24.62) | 44.73 (15.16) | Ns | Ns | Ns |
| Surprise | 43.59 (18.85) | 31.09 (15.84) | 30.72 (26.42) | 38.19 (15.46) | Ns | Ns | Ns |
| Total | 43.06 (15.65) | 32.95 (16.58) | 30.86 (24.94) | 39.40 (14.11) | Ns | Ns | Ns |

*based on ANOVA test
**based on post hoc group comparisons
*** based on post hoc group comparisons adjusted for age and sex

**Table S5.** Dwell time within Areas of Interest (AOI) Mouth in Ekman faces across groups

|  | bvAD | tAD | bvFTD | CN | p-value* | Group differences, unadjusted | Group differences, adjusted ** |
| --- | --- | --- | --- | --- | --- | --- | --- |
| N_AOI_ | 12 | 13 | 12 | 16 |  |  |  |
| Happy | 21.10 (12.43) | 34.47 (13.19) | 20.40 (14.46) | 34.55 (7.73) | Ns | bvAD<CN, p=0.005, bvFTD<CN, p=0.003, bvAD<tAD, p=0.007, bvFTD<tAD, p=0.005 | bvAD<CN, p=0.009, bvFTD<CN, p=0.005, bvAD<tAD, p=0.006, bvFTD<tAD, p=0.003 |
| Angry | 18.55 (9.27) | 33.17 (11.17) | 19.60 (15.86) | 30.97 (11.82) | Ns | bvAD<CN, p=0.01, bvFTD<CN, p=0.02, bvAD<tAD, p=0.004, bvFTD<tAD, p=0.008 | bvAD<CN, p=0.006, bvFTD<CN, p=0.01, bvAD<tAD, p=0.002, bvFTD<tAD, p=0.004 |
| Disgust | 19.69 (10.02) | 33.93 (13.42) | 20.18 (15.04) | 31.71 (13.24) | Ns | Ns | bvAD<CN, p=0.01, bvFTD<CN, p=0.02, bvAD<tAD, p=0.002, bvFTD<tAD, p=0.004 |
| Fear | 19.30 (9.82) | 32.67 (12.84) | 18.89 (13.75) | 30.15 (11.53) | Ns | bvAD<CN, p=0.02, bvFTD<CN, p=0.02, bvAD<tAD, p=0.008, bvFTD<tAD, p=0.006 | bvAD<CN, p=0.01, bvFTD<CN, p=0.01, bvAD<tAD, p=0.003, bvFTD<tAD, p=0.003 |
| Sad | 15.29 (8.44) | 30.27 (12.08) | 17.78 (15.52) | 26.69 (12.51) | Ns | bvAD<CN, p=0.02, bvAD<tAD, p=0.004, bvFTD<tAD, p=0.02 | bvAD<CN, p=0.01, bvFTD<CN, p=0.01, bvAD<tAD, p=0.0006, bvFTD<tAD, p=0.005 |
| Surprise | 18.49 (10.46) | 31.81 (12.58) | 19.33 (13.56) | 28.15 (10.66) | Ns | bvAD<CN, p=0.04, bvAD<tAD, p=0.007, bvFTD<tAD, p=0.01 | bvAD<CN, p=0.04,  bvAD<tAD, p=0.004,  bvFTD<tAD, p=0.007 |
| Total | 18.74 (9.53) | 32.72 (12.08) | 19.37 (14.28) | 30.37 (10.57) | Ns | bvAD<CN, p=0.01, bvFTD<CN, p=0.02, bvAD<tAD, p=0.004, bvFTD<tAD, p=0.006 | bvAD<CN, p=0.008, bvFTD<CN, p=0.01, bvAD<tAD, p=0.001,  bvFTD<tAD, p=0.003 |

*based on ANOVA test
**based on post hoc group comparisons
*** based on post hoc group comparisons adjusted for age and sex

**Level 2 Empathy- Interpersonal Reactivity Index**

**Table S6.** Scores on the Interpersonal Reactivity Index, a questionnaire measuring empathy, across diagnostic groups

|  | bvAD | tAD | bvFTD | CN | p-value* | Group differences, unadjusted | Group differences, age and sex adjusted** |
| --- | --- | --- | --- | --- | --- | --- | --- |
| n | 9 | 10 | 11 | 16 |  |  |  |
| Perspective Taking | 11.33 (4.58) | 13.00 (3.19) | 8.73 (5.61) | 15.50 (3.35) | 0.0008 | bvAD<CN, p=0.02, bvFTD<CN, p=0.0002, bvFTD<tAD, p=0.02 | bvAD<CN, p=0.04,  bvFTD<CN, p=0.0004, bvFTD<tAD, p=0.03 |
| Empathetic Concern | 12.89 (4.28) | 12.40 (4.19) | 12.00 (3.00) | 13.69 (1.14) | 0.15 | Ns | Ns |
| Fantasy | 10.33 (5.22) | 13.00 (3.09) | 8.64 (3.96) | 13.63 (1.75) | 0.006 | bvAD<CN, p=0.03, bvFTD<CN, p=0.0007, bvFTD<tAD, p=0.006 | bvAD<CN, p=0.008,  bvFTD<CN, p=0.004, bvFTD<tAD, p=0.01 |
| Personal Distress | 13.11 (3.52) | 14.50 (2.95) | 14.45 (3.01) | 14.31 (1.96) | 0.74 | Ns | Ns |

*based on ANOVA test
**based on post hoc group comparisons
*** based on post hoc group comparisons adjusted for age and sex

**Level 2 Empathy – empathy eliciting videos**

**Table S7.** Results of empathy videos across diagnostic groups: empathetic concern and personal distress scores and galvanic skin response while watching empathy eliciting videos

|  | bvAD | tAD | bvFTD | CN | p-value* | Group differences, unadjusted | Group differences, age and sex adjusted** |
| --- | --- | --- | --- | --- | --- | --- | --- |
| n | 11 | 11 | 11 | 15 |  |  |  |
| Empathetic concern | 2.55 (1.08) | 2.61 (1.21) | 2.11 (1.20) | 2.82 (0.68) | 0.38 | Ns | Ns |
| Personal distress | 1.61 (0.66) | 1.64 (0.66) | 1.87 (1.20) | 1.63 (0.52) | 0.87 | Ns | Ns |
| n | 11 | 11 | 10 | 15 |  |  |  |
| GSR PPM uplifting video | 1.56 (1.99) | 1.56 (1.99) | 2.31 (2.66) | 2.14 (1.84) | 0.28 | Ns | Ns |
| GSR PPM distressing video | 1.97 (2.41) | 1.97 (2.41) | 2.06 (3.10) | 1.58 (1.74) | 0.56 | Ns | Ns |

*based on ANOVA test
**based on post hoc group comparisons
*** based on post hoc group comparisons adjusted for age and sex

**Level 3 Social Norms Questionnaire**

**Table S8**. Scores on Social Norms Questionnaire across diagnostic groups

|  | bvAD | tAD | bvFTD | CN | p-value* | Group differences, unadjusted** | Group differences, sex and age adjusted*** |
| --- | --- | --- | --- | --- | --- | --- | --- |
| n | 11 | 11 | 11 | 15 |  |  |  |
| SNQ total score | 16.00 (1.61) | 17.82 (2.14) | 15.18 (2.23) | 18.13 (1.30) | 0.0004 | bvAD<CN, p=0.005, bvFTD<CN, p=0.0002, bvAD<tAD, p=0.02, bvFTD<tAD, p=0.001 | bvAD<CN, p=0.02, bvFTD<CN, p=0.0003, bvAD<tAD, p=0.04, bvFTD<tAD, p=0.001 |
| SNQ break score | 2.09 (1.51) | 1.09 (0.54) | 1.73 (1.42) | 1.40 (0.99) | 0.24 | Ns | Ns |
| SNQ overadherence score | 3.91 (1.58) | 3.09 (2.12) | 5.09 (2.30) | 2.47 (1.06) | 0.005 | bvAD<CN, p=0.05, bvFTD<CN, p=0.0006, bvFTD<tAD, p=0.01 | bvFTD<CN, p=0.001, bvFTD<tAD, p=0.01 |

*based on ANOVA test
**based on post hoc group comparisons
*** based on post hoc group comparisons adjusted for age and sex

**Level 3 Moral dilemmas**

**Table S9.** Scores on and galvanic skin response to moral dilemmas, across diagnostic groups

|  | bvAD | tAD | bvFTD | CN | p-value* | Group differences, unadjusted | Group differences, age and sex adjusted** |
| --- | --- | --- | --- | --- | --- | --- | --- |
| n | 10 | 11 | 11 | 15 |  |  |  |
| Personal dilemma, percentage rational responses | 50.00 | 36.36 | 18.18 | 20.00 | 0.82 | Ns | Ns |
| Impersonal dilemma, percentage rational responses | 90.00 | 90.91 | 81.82 | 93.33 | 0.31 | Ns | ns |
| n | 10 | 11 | 9 | 13 |  |  |  |
| Impersonal dilemma, GSR PPM | 3.17 (2.34) | 3.16 (3.65) | 1.48 (1.71) | 2.28 (2.01) | 0.42 | Ns | Ns |
| Personal dilemma, GSR PPM | 2.89 (2.29) | 3.43 (4.07) | 0.86 (1.08) | 3.16 (3.32) | 0.22 | Ns | bvFTD<CN, p=0.02 |

*based on ANOVA test
**based on post hoc group comparisons
*** based on post hoc group comparisons adjusted for age and sex

**Figure S1.** Example of Area of Interest imposed on the Ekman faces to retrieve eyetracking dwell time

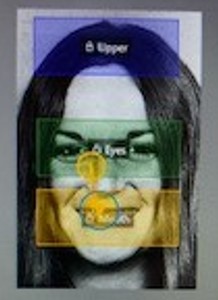

**Figure S2.** Matrices of missing values on the Ekman and SNQ prior to imputing procedures.

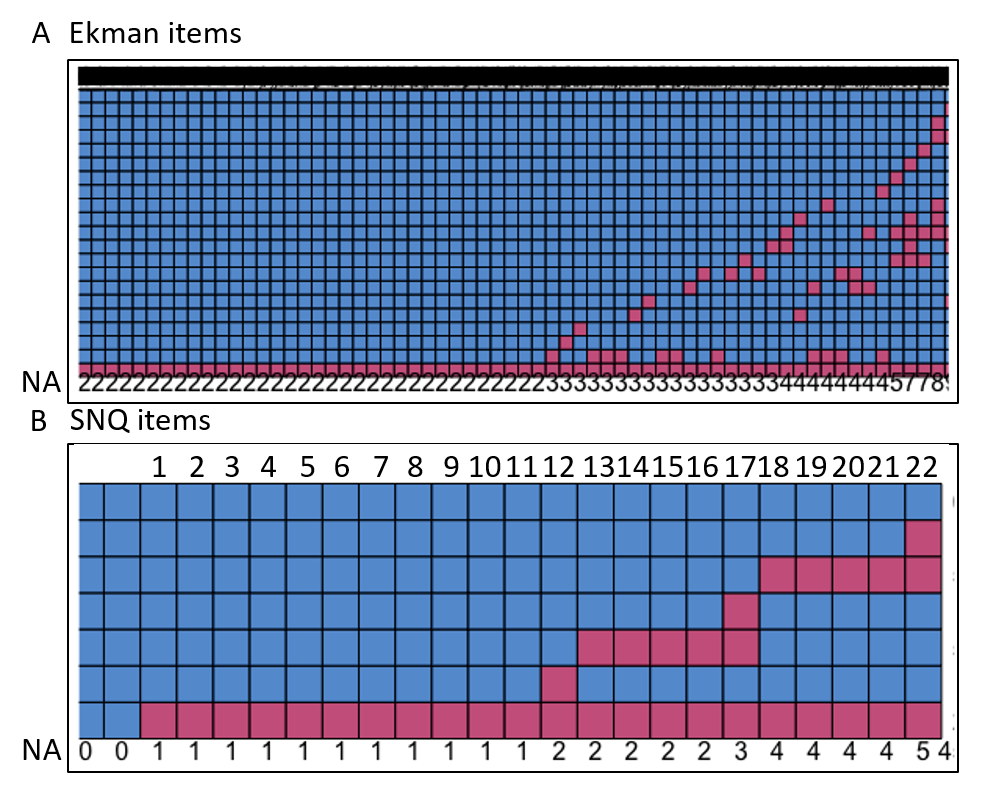

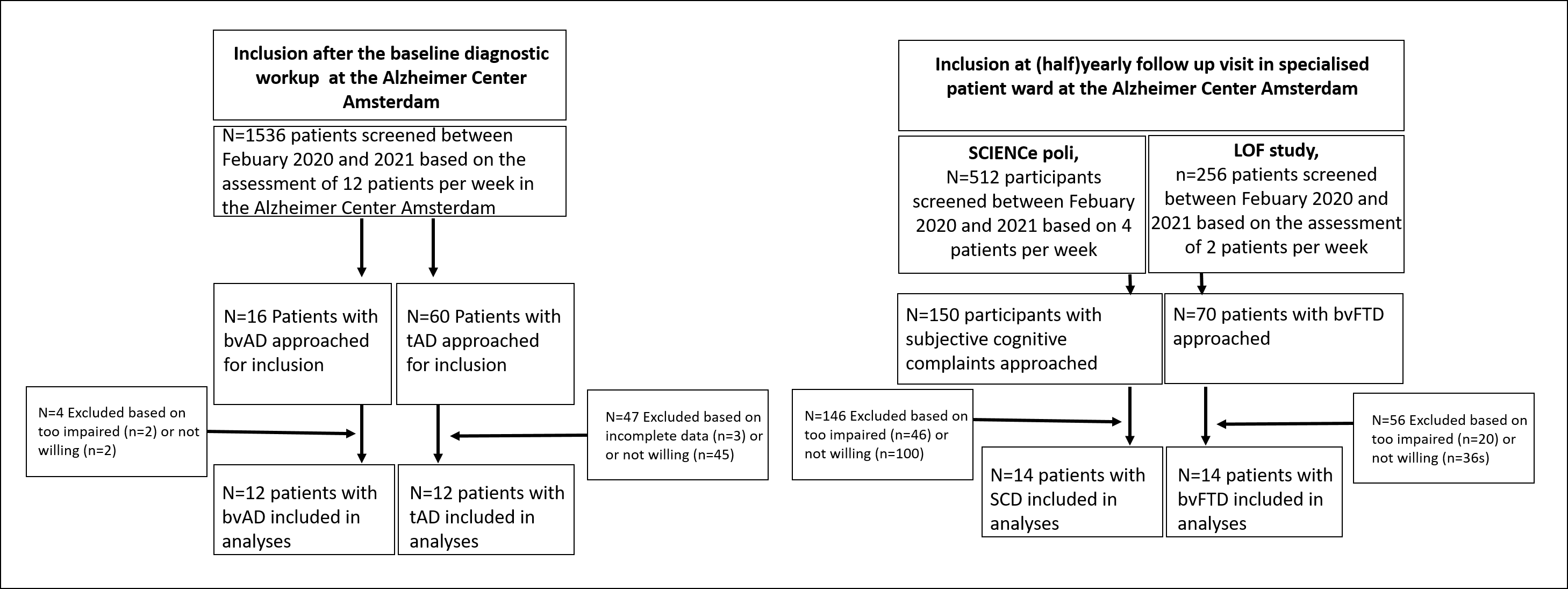
**Figure S3**. Inclusion flow of participants into the current study
